# Safe attitudes, uncertain implementation: concussion knowledge, attitudes, and confidence among parents and staff in junior community Australian football

**DOI:** 10.64898/2026.07.20.26358513

**Authors:** Hunter Bennett, Chloe Fletcher, Elizabeth Jemson-Ledger, Joel Garrett

**Affiliations:** Allied Health and Human Performance, Adelaide University, Adelaide, Australia; School of Alliance for Research in Exercise, Nutrition, and Activity (ARENA), Adelaide, Australia; Adelaide University Rural Health, Adelaide University, Adelaide, Australia; IIMPACT in Health, Adelaide University, Adelaide, Australia; Advanced Neuro Rehab, Adelaide, Australia; School of Allied Health, Sport and Social Work, Griffith University, Gold Coast, Australia; Australian Centre for Precision Health and Technology (PRECISE), Griffith University, Gold Coast Australia

## Abstract

**Introduction:** Parents and club staff are primarily responsible for recognising and managing concussion in junior community Australian Football (ARF), yet no recent research has examined their knowledge or practices. This study assessed concussion knowledge, attitudes, and confidence among parents and staff in junior (under-16 years) community ARF, and identified management practices and perceived barriers.

**Methods:** An online survey assessing concussion knowledge and attitudes (RoCKAS-ST), confidence, and AFL-specific guideline knowledge was distributed nationally to parents and staff of junior community ARF players. A subsample whose child had a concussion in the previous 12 months completed additional questions on management practices. Closed-ended responses were analysed descriptively and using Pearson’s correlation. Open-ended responses underwent qualitative content analysis.

**Results:** Responses (n=223) reflected high general concussion knowledge (76.6% of maximum) and positive attitudes toward safe concussion management (86.8% of maximum), but AFL-specific guideline knowledge was limited (49.8%). Self-reported confidence was not associated with knowledge (r = -.04) or attitudes (r = .09). Among 58 respondents whose child recently sustained a concussion, medical evaluation (74.1%) and clearance prior to return (60.3%) were common, but graded return-to-play protocol use (50.0%) and club support during recovery (43.1%) were inconsistent. Knowledge gaps (70.4%), cultural attitudes favouring playing through symptoms (29.6%), and inadequate volunteer capacity (29.6%) were most common barriers.

**Conclusions:** Parents and staff in junior community ARF hold safe attitudes toward concussion but lack the sport-specific knowledge, structural support, and resources to translate these attitudes into practice. Targeted, mandated education, and improved support are needed to close this gap.

## 1. Introduction

The 6th International Conference on Concussion in Sport defines concussion as a traumatic brain injury caused by a direct blow to the head, neck, or body resulting in an impulsive force being transmitted to the brain, initiating a neurotransmitter and metabolic cascade with possible axonal injury, blood flow change, and inflammation.^1^ Although most individuals recover fully, the high prevalence of concussion and the persistence of a range of clinical symptoms contribute to a substantial ongoing burden on healthcare resources.^2^

Children represent a population of particular concern.^3^ Research has indicated that children sustaining a concussion are at risk of various long-term deficits, including reductions in school attendance, potential declines in academic performance,^4, 5^ mental health difficulties,^6^ and behavioural disturbances.^7^ Of additional concern is the elevated risk of re-injury following an initial concussion. Recent meta-analytic research has shown that the risk of sustaining a concussion is more than three times greater in children with a previous concussion compared with those without a prior concussion.^8^ This vulnerability to repeat injury may carry serious clinical implications, as premature return to play has been proposed to expose young athletes to the risk of second impact syndrome, a rare but potentially catastrophic condition in which a second concussion sustained before complete recovery from the first can result in rapid, diffuse cerebral oedema.^9^

Sport is a leading context in which concussions occur in children and adolescents. Sport-related concussions (SRC) are a concern for young athletes due to their potential for long-term health problems.^3^ Incidence rates vary across sports, with collision and contact sports carrying the highest risk.^10^ In Australia, Australian football (ARF) is one of the most widely played team sports, representing the second most popular team sport in the country.^11^ ARF is a collision sport that involves significant player-to-player contact, and SRC is among the more common injuries in elite, sub-elite, and community settings.^12–14^ In community junior ARF specifically, the head has been identified as the most frequently reported injured body region, with approximately one in every two reported head injuries resulting in a diagnosed or suspected SRC.^15^

In 2023, approximately 300,000 children under 16 years of age participated in community ARF across Australia, with participation increasing yearly. For child athletes, the responsibility for SRC recognition and management falls primarily on the adults around them (i.e., parents and club staff), rather than on the athletes themselves. Parents and coaches play key roles in preventing, recognising, and responding to SRC in youth and high school sports.^16^ However, while coaches and parents can often identify common physical signs and symptoms of SRC, such as headache and dizziness, they tend to show considerably less awareness of emotional symptoms and of appropriate management and return-to-play protocols.^17^ Critically, people in sport have shown measurable improvements in knowledge and changes in attitudes following targeted concussion education programs.^18^ This highlights the potential for targeted education interventions to improve SRC awareness and management in community ARF. However, what aspects of awareness and management these interventions should target remains unclear.

To date, three studies have explored the concussion knowledge and management practices of parents and staff involved in junior ARF. Haran et al. (2016) found that, concerningly, 42% of concussed children were not managed according to recommended guidelines, and most parents (93%) and athletes (96%) were unaware of the recommendations made by return-to-play protocols.^19^ Similarly, Hecimovich et al. (2016) assessed concussion knowledge in 1,441 parents and 284 male youth ARF players, revealing large variations in knowledge across participants. Notably, parents who had received prior concussion education demonstrated significantly greater awareness, further highlighting the potential value and importance of education programs in this population.^20^ Expanding on this, White et al. (2017) explored how parents of community-level ARF players (under 16) perceived and experienced concussion guidelines. A key finding was that parents wanted more accessible information on concussion protocols and return-to-play procedures.^21^

Despite these contributions, significant gaps remain. It has been more than 10 years since any research has examined concussion knowledge and management practices in junior ARF. This is particularly noteworthy as the AFL introduced updated community concussion guidelines in 2024 (which include the recommendation for each club to have a dedicated ‘concussion officer’),^22^ and there has been substantial growth in junior female participation since the inception of the Australian Football Women’s League (AFLW) in 2017.^23^ Given the high and increasing rate of participation in junior community ARF, the elevated SRC risk inherent to the sport, and the critical role of parents and club staff in safeguarding child athletes, contemporary research into concussion knowledge and management practices is needed.

This exploratory study therefore aimed to: (1) assess the knowledge, attitudes, and confidence of club staff and parents in recognising, managing, and supporting concussion in junior (under 16 years) community ARF, and (2) identify current concussion management practices, including adherence to return-to-play protocols and barriers to proper rehabilitation, in this population.

## 2. Methods

### 2.1 Experimental approach to the problem

A purpose-built survey developed using Qualtrics (Qualtrics, Provo, UT) was distributed to parents and staff of junior ARF (<16 years) players to assess their knowledge, attitudes, and confidence of concussion and current concussion guidelines (Part A, described below). A subset of participants completed additional questions pertaining to current concussion management practices (Part B). The survey was distributed widely across Australia using a variety of communication channels, including community sporting clubs, social media, and personal networks. Prior to distribution, the survey was evaluated for content-accuracy by a qualified Neurological Physiotherapist and piloted by five individuals involved in junior (<16 years) ARF (i.e., parents and coaches) to ensure readability and acceptability amongst the target audience. This study was pre-registered on the Open Science Framework (OSF) (https://osf.io/knqw6/overview), and the final survey can be accessed via the OSF project linked to this study (https://osf.io/g5x6r). This study was approved by the University of South Australia Human Research Ethics Committee (project number 207114).

### 2.2 Participants

Assuming a total population of approximately 300,000 children <16 years playing ARF, using an estimated proportion of 0.5, a 95% confidence level, and a ±5% margin of error, we aimed to recruit 384 participants. The survey was closed after 408 had commenced the survey. However, of these, 82 (20.1%) provided insufficient data to be included. A further 103 (25.2%) were removed due to having an extremely high likelihood of being automated and/or AI generated. After data cleaning, 223 responses were included in the analysis. Recruitment ceased after available recruitment pathways were exhausted and recruitment slowed substantially. With the final sample of 223 participants, using the same parameters (95% confidence level, estimated proportion of 0.5), this corresponds to a margin of error of ±6.56%. Informed consent was obtained from each subject who read the information sheet detailing the research aims before starting the survey. All individuals aged >18 years involved with junior (<16 years) community ARF as staff, or parents of junior (<16 years) community ARF players, were eligible to participate.

### 2.3 Outcome measures, Part A

All participants eligible for inclusion completed surveys pertaining to the following categories.

- **Concussion knowledge and attitudes**: were assessed using the Rosenbaum concussion knowledge and attitudes survey (RoCKAS-ST).^24^
- **Concussion confidence**: was assessed using Likert type responses (1 = Not at all confident, 5 = Extremely confident) to statements around confidence recognizing, knowing what to do after, supporting recovery from, and deciding when to return to sport after, a concussion.
- **Sport specific concussion guideline knowledge**: was assessed via purpose-built questions based upon the Australian Football League 2024 community concussion guidelines.^22^
- **Concerns, barriers, and resource needs**: were identified via open-ended questions pertaining to these three factors.

### 2.4 Outcome measures, Part B

A subset of participants whose child had experienced a concussion in the 12 months prior to them completing the study completed an additional survey pertaining to:

- **Concussion management:** was assessed via purpose-built questions pertaining to aspects of concussion management, including; what occurred post-concussion, who diagnosed the concussion, time missed from school/sport as a result of the concussion, symptoms experienced, and return-to-play process after the concussion.
- **Concussion management experiences and needs:** were identified via open-ended questions pertaining to what worked well during the concussion recovery process, what challenges were faced during the recovery process, and what resources would have helped during the recovery process.

### 2.5 Data preparation and analysis

Survey data was exported from Qualtrics to an Excel spreadsheet, where 408 responses were screened and partial or bot- or AI-generated responses were removed. Responses were assumed to be bot- or AI-generated if they demonstrated repeated identifiers across submissions (i.e., the same IP address multiple times with the exact same responses), implausible geographic origin relative to the target population, and/or systematic patterns of non-substantive open-ended responses (e.g., numeric or irrelevant entries), suggestive of automated or coordinated response generation.

Responses were also excluded if they did not answer enough questions to contribute to any part of the survey (e.g., only provided demographic data). Responses to “textbox” style questions were assessed and manually edited to ensure formatting constancy (i.e., conversion of years to months, or weeks to days) when required.

For both Part A and B of the survey, closed-ended responses were reported descriptively using frequencies and percentages, means and standard deviations, and median and interquartile ranges. Associations between survey items were explored using Pearsons’s correlation coefficients.

Quantitative synthesis was conducted using R studio (version 4.5.2). Responses to open-ended questions were analysed separately using qualitative content analysis. Content analysis was selected given the nature of the data (i.e., short free-text responses to open-ended survey questions) and its compatibility with a descriptive coding approach and quantification of code frequency. A bottom-up, inductive approach was used, whereby responses were read in full, and codes were generated directly from their content rather than from a predetermined framework. Codes with similar content were subsequently grouped into categories within each question. Frequencies are reported as the number and percentage of substantive responses assigned to each category, and responses addressing more than one category were coded to all relevant categories. As a result, frequencies within a given question can sum to more than 100%. Responses that were non-substantive (e.g., “not sure”) were identified and excluded. For transparency, all de-identified quantitative and qualitative data are available on the Open Science Framework project linked to this study (https://osf.io/cwdu4/files/osfstorage).

## 3. Results

### 3.1 Participants

A total of 408 individuals commenced the survey. Of these, 82 (20.1%) provided insufficient data to be included. A further 103 (25.2%) were removed due to having an extremely high likelihood of being automated and/or AI-generated. This left a total of 223 individuals involved in junior (<16 years) community Australian Rules Football (ARF) who completed the survey. The majority identified as parents (n = 193, 86.5%), with some identifying as trainers or first aid officers (n = 25, 11.2%), club administrators (n = 18, 8.1%), team managers (n = 17, 7.6%), and coaches (n = 14, 6.3%), or in other roles (n = 10, 4.5%). Because respondents could select multiple roles, these percentages sum to greater than 100%. With respect to prior club involvement, 23.3% (n = 52) had served as team managers, 21.5% (n = 48) as first aid officers or trainers, 20.6% (n = 46) as committee members, 9% (n = 20) as coaches, and 12.1% (n = 27) in other roles (e.g., runners, assistant coaches, umpires). Of the 223 respondents, 215 (96.4%) provided a postcode from which their broad geographical location could be reported (Table 1).

**Table 1:**
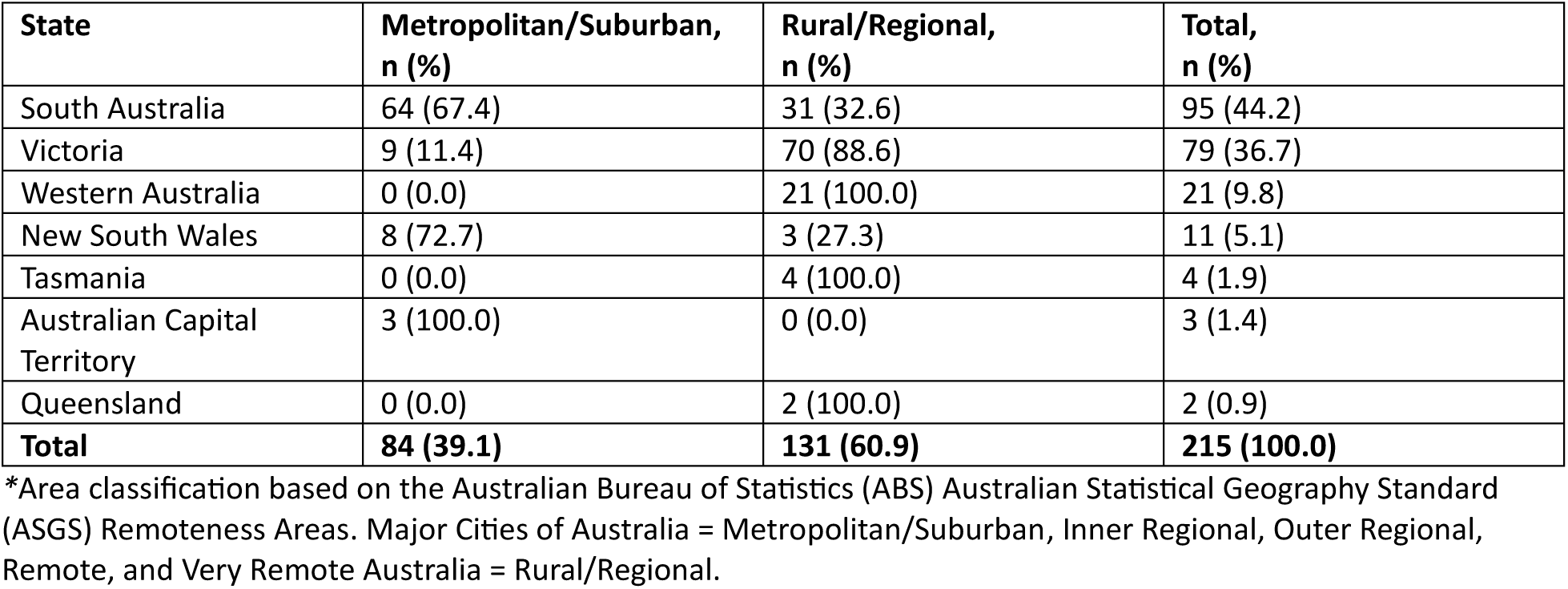
Geographic distribution of respondents by state and area classification (n = 215).

| State | Metropolitan/Suburban, n (%) | Rural/Regional, n (%) | Total, n (%) |
| --- | --- | --- | --- |
| South Australia | 64 (67.4) | 31 (32.6) | 95 (44.2) |
| Victoria | 9 (11.4) | 70 (88.6) | 79 (36.7) |
| Western Australia | 0 (0.0) | 21 (100.0) | 21 (9.8) |
| New South Wales | 8 (72.7) | 3 (27.3) | 11 (5.1) |
| Tasmania | 0 (0.0) | 4 (100.0) | 4 (1.9) |
| Australian Capital Territory | 3 (100.0) | 0 (0.0) | 3 (1.4) |
| Queensland | 0 (0.0) | 2 (100.0) | 2 (0.9) |
| **Total** | **84 (39.1)** | **131 (60.9)** | **215 (100.0)** |

\*Area classification based on the Australian Bureau of Statistics (ABS) Australian Statistical Geography Standard (ASGS) Remoteness Areas. Major Cities of Australia = Metropolitan/Suburban, Inner Regional, Outer Regional, Remote, and Very Remote Australia = Rural/Regional.

Approximately 8% of respondents identified as Aboriginal and/or Torres Strait Islander (Aboriginal: n = 17, 7.6%; Torres Strait Islander: n = 1, 0.4%), with 7 respondents (3.1%) not providing a response to this question. The median number of children per respondent was 2 (mean [M] = 2.5, standard deviation [SD] = 0.9, range: 1-7), and the median number of junior ARF players they were a parent of was 1 (M = 1.6, SD = 0.7, range: 0-4). All demographic data are presented in Table 2.

**Table 2:**
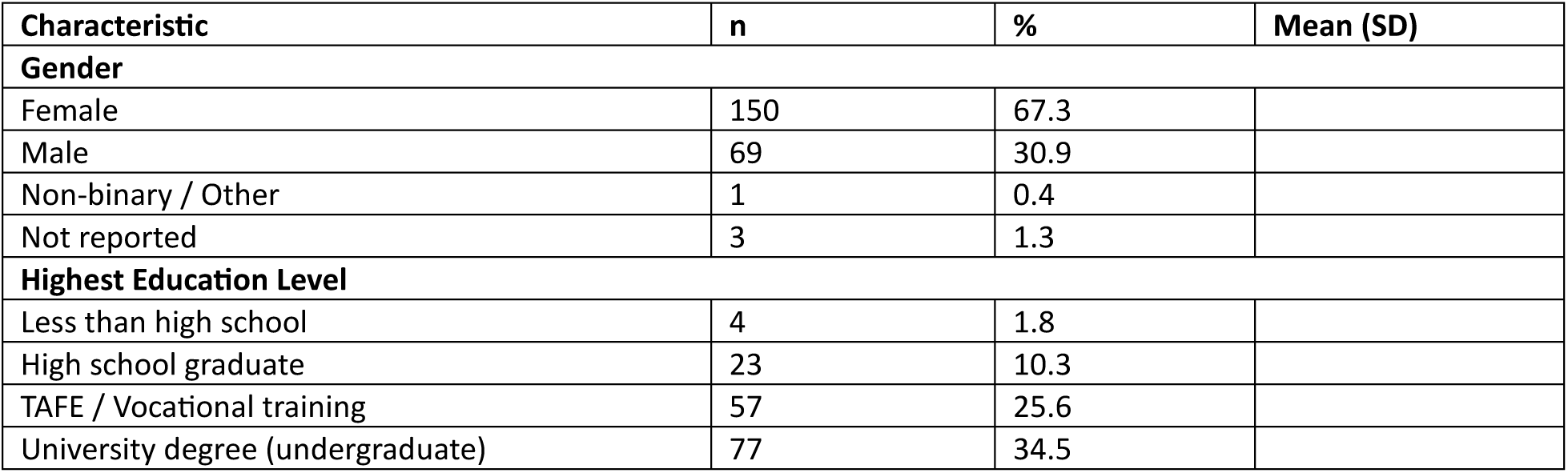
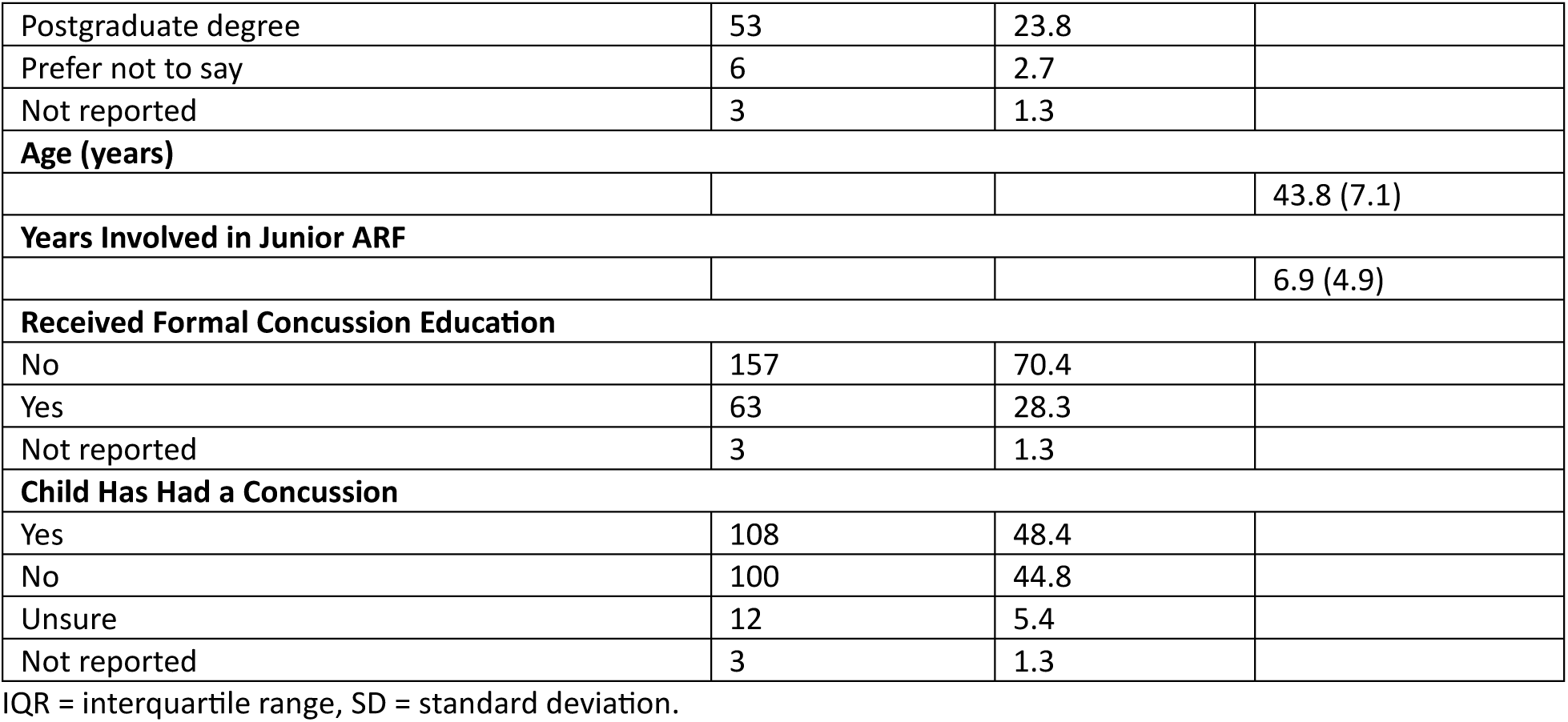
Demographic information (n=223)

### 3.2 Part A, Quantitative Data

#### 3.2.1 Concussion Knowledge Index

The Concussion Knowledge Index (CKI; true-false responses; maximum possible score = 25) assesses general concussion knowledge across three sections of the RoCKAS-ST (section 1, 2, and 5). Across 205 respondents with complete CKI data, the average CKI score was 19.15 (SD = 2.65, range: 6-23), corresponding to 76.6% of the maximum possible score and reflecting accurate knowledge. The distribution was negatively skewed (skew = −2.29), indicating that most respondents performed above the midpoint. Descriptive statistics for all scales and subscales are presented in Table 3. Internal consistency for CKI was acceptable (Cronbach’s α = .69).

**Table 3:**
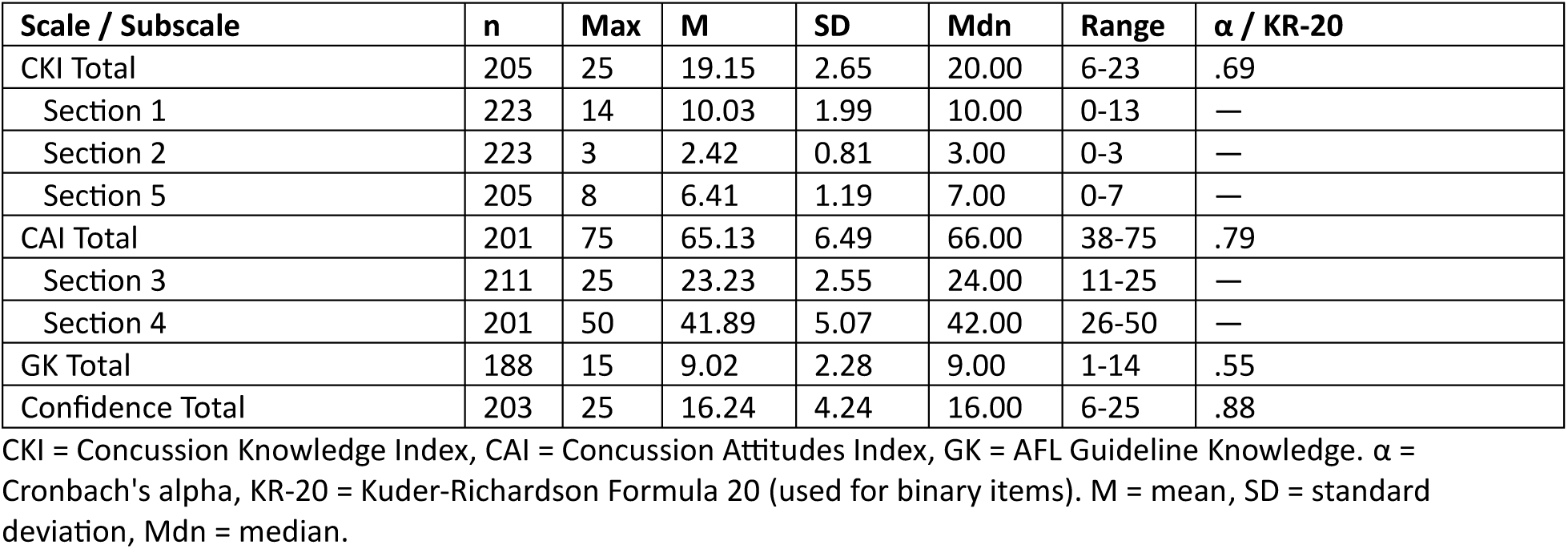
Concussion knowledge, attitudes, guidelines knowledge, and confidence results.

At the item level, the lowest rates of correct responses were observed for Item 11, which concerns whether concussion typically results in visible abnormalities on brain imaging, and Item 18, which addresses the risk of long-term harm associated with multiple concussions. These findings suggest that misconceptions persist in these key areas.

#### 3.2.2 Concussion AFtudes Index

The Concussion Attitudes Index (CAI; five-point Likert scale from strongly disagree to strongly agree; maximum possible score = 75) measures attitudes toward safe concussion management across two sections (section 3 and 4), with higher scores reflecting safer attitudes. Internal consistency for the CAI was good (Cronbach’s α = .79). Among the 201 respondents with complete CAI data, the mean total CAI score was 65.13 (SD = 6.49, range: 38-75), corresponding to 86.8% of the maximum possible score. The distribution was negatively skewed (skew = −0.97), indicating that most respondents endorsed generally safe attitudes toward concussion management.

#### 3.2.3 AFL Guideline Knowledge

The AFL Guideline Knowledge scale (GK; mix of multiple choice and true/false questions; maximum possible score = 15) assessed participants’ familiarity with the AFL Community Concussion Guidelines (2024), using purpose-built items. Internal consistency was acceptable (KR-20 = .55). 192 respondents provided data for this aspect of the survey (188 answered every question), and the mean GK score was 9.02 (SD = 2.28, range: 1-14), representing 60.0% of the maximum possible score. The distribution was negatively skewed (skew = −0.27), indicating that many respondents performed above the midpoint, though considerable variability was observed.

At the item level, respondents performed well on items concerning who can diagnose a concussion and provide clearance for return to play (89.6% of respondents correctly identified a medical doctor should assess a player to diagnose a suspected concussion; 95.3% correctly identified that trainers and sports staff can override a doctor’s clearance if they think a player is ready), with three quarters (75.5%) correctly identifying that a child cannot return to play until symptoms have resolved and they have received a medical clearance. Conversely, performance was poor on items relating to the minimum monitoring period following a suspected concussion, with most respondents (10.6% were correct) failing to identify the guideline-specified response of ‘At least 3 hours’ and instead often selecting ‘Until symptoms resolve’. Respondents also frequently incorrectly identified appropriate return to play timelines, where only 16.8% correctly identified 7 days as the minimum amount of time a player must be exposed to sport specific non-contact and contact training before returning to competition, although more than half (54.2%) correctly identified 21 days as the earliest day a player may return to competitive football after a concussion.

#### 3.2.4 Confidence in Concussion Management

Self-reported confidence in concussion management was assessed across five questions using a five-point Likert scale (1 = Not at all confident, 2 = Slightly confident, 3 = Moderately confident, 4 = Very confident, 5 = Extremely confident). Internal consistency was excellent (Cronbach’s α = .88). Among the 203 respondents with complete confidence data, the mean total confidence score was 16.24 (SD = 4.24, range: 6-25), corresponding to a mean item rating of 3.25 (*SD* = 0.85), indicative of moderate confidence overall.

At the question level, respondents reported highest confidence in knowing where to take their child for assessment, followed by supporting a child through recovery, knowing what to do if a concussion is suspected, deciding when it is safe to return to training or play, and recognising the signs and symptoms of concussion.

#### 3.2.5 Relationships Between Scales

CKI was moderately correlated with both CAI (r = .39) and GK (r = .37), and CAI was also moderately correlated with GK (r = .24) (all p <0.001), suggesting that respondents with greater general concussion knowledge tend to hold safer attitudes and demonstrate better familiarity with AFL-specific guidelines. Conversely, self-reported confidence was not meaningfully associated with general concussion knowledge or attitudes, whereby correlations with CKI (r = -.04) and CAI (r = .09) were near zero and non-significant (both p > .18). The only significant association involving confidence was with GK (r = .33, p < .001), indicating a modest relationship between familiarity with AFL-specific guidelines and confidence in concussion management.

### 3.3 Part A, Qualitative Data

#### 3.3.1 Concerns about concussion in Junior ARF

Respondents were asked to identify their biggest concerns about concussion in junior Australian Rules Football. Of 179 responses received, 175 provided information that could be coded (four excluded as non-substantive). Seven categories were identified. **Long-term brain health and the effects of repeated concussions** (C1) was the most common concern (n = 72, 41.1%). Respondents expressed worry about cumulative neurological, cognitive, emotional, and academic consequences of concussion on developing brains, including depression, learning difficulties, and irreversible damage. Several connected their concerns to their child’s health across the lifespan. For example, *”that my child over time will get a brain injury that will affect him into his adult life”* and *”that it is something that can occur at junior level but may have lasting effects long down the line in their lives.”* The next most frequently cited categories were **lack of education and awareness** (C5; n = 49, 28.0%) and **inadequate recognition and diagnosis** (C4; n = 44, 25.1%). C5 captured concerns about systemic attitude and knowledge deficits across the football community, including the view that *”as a community, we are in the infancy of understanding the seriousness of concussion.”* Conversely, C4 captured more specific concerns about failure to identify concussion at the time of injury, including undiagnosed mild concussions, delayed symptom onset. Notably, a small number of respondents raised the risk of over-diagnosis or overly cautious removal from play as concerns.

The remaining four categories were each identified by one fifth of respondents or fewer. **Players hiding or underreporting symptoms** (C2; n = 35, 20.0%) reflected concern about the pressures that may lead young athletes to conceal symptoms to remain on the field. For example, *“it happens so easily. My boys want to play very badly, and they know that admitting to concussion symptoms may result in them not playing, so they may not tell me.”* **Game rules, umpiring and physical risk factors** (C7; n = 34, 19.4%) encompassed concerns about dangerous tackling that goes unpenalized, player size, developmental mismatches, and the absence of teaching around safe play. For example, *”the same players are sling tackling and doing high tackles but aren’t sent off the field.”* **Volunteer and structural capacity at club level** (C6; n = 32, 18.3%) captured concerns about the absence of clear procedures and qualified personnel. This included the inability of on-field staff to make decisions under pressure from coaches and families, with one respondent noting *”medics not being able to make the decision of when to pull out the players due to pressure from the player, family, or coach.”* Finally, **premature return to play** (C3; n = 22, 12.6%) was the least common category, though its relatively low frequency should be interpreted cautiously given that concerns about return to play were often embedded within other responses. All categories identified across open-ended questions in Part A are presented in Table 4.

**Table 4:** Summary of Part A qualitative analysis results.

| Category | n | % |
| --- | --- | --- |
| **Part A: All respondents** |  |  |
| **Biggest concerns about concussion in junior ARF (n = 175 substantive responses)** |  |  |
| Long-term brain health and the effects of repeated concussions (C1) | 72 | 41.1% |
| Lack of education and awareness (C5) | 49 | 28.0% |
| Inadequate recognition and diagnosis (C4) | 44 | 25.1% |
| Players hiding or underreporting symptoms (C2) | 35 | 20.0% |
| Game rules, umpiring, and physical risk factors (C7) | 34 | 19.4% |
| Volunteer and structural capacity at club level (C6) | 32 | 18.3% |
| Premature return to play (C3) | 22 | 12.6% |
| **Perceived barriers to proper concussion management (n = 159 substantive responses)** |  |  |
| Knowledge and education gaps (B1) | 112 | 70.4% |
| Cultural attitudes and social pressure to play on (B2) | 47 | 29.6% |
| Inadequate volunteer capacity and untrained staff (B3) | 47 | 29.6% |
| Inconsistent protocols and lack of enforcement (B5) | 29 | 18.2% |
| Limited access to qualified medical professionals (B4) | 25 | 15.7% |
| Financial and resource constraints (B6) | 18 | 11.3% |
| Player underreporting and symptom concealment (B7) | 11 | 6.9% |
| **Desired knowledge about concussion management (n = 121 substantive responses)** |  |  |
| Education, training, and club-level resources (M5) | 47 | 38.8% |
| Recognising symptoms and early identification (M2) | 46 | 38.0% |
| Return-to-play protocols and recovery timelines (M1) | 31 | 25.6% |
| Home management and practical post-incident guidance (M4) | 31 | 25.6% |
| Long-term effects and ongoing monitoring (M3) | 29 | 24.0% |
| Equipment, technology, and prevention (M6) | 19 | 15.7% |

#### 3.3.2 Perceived Barriers to Concussion Management

Respondents identified seven barriers to proper concussion management across 159 responses (12 excluded as non-substantive). The most common barrier was **knowledge and education gaps** (B1; n = 112, 70.4%), reflecting a concern that insufficient understanding of concussion symptoms, risks, and management protocols, among coaches, parents, players and staff remains a key obstacle. As one respondent summarised, *”Education. Not all of the trainers in junior community sport have concussion training.”* **Cultural attitudes and social pressure to play on** (B2) and **inadequate volunteer capacity and untrained staff** (B3) were the next most frequently identified barriers (both n = 47, 29.6%). The former captured persistent “old school” and “she’ll be right” mentalities. For example, a respondent described: *”old school mentality still exists where it’ll be fine attitude and have a couple of days off and play next week, not till all symptoms have subsided.”* The latter reflected concerns about the reliance on rotating, untrained parent volunteers as the primary first-aid presence at games, with one respondent stating *”Relies on volunteers. But what qualifications do first aiders have in junior footy?”*

The remaining four barriers were identified less frequently but remained substantive. **Inconsistent protocols and lack of enforcement** (B5; n = 29, 18.2%) captured variation in concussion management across clubs and governing bodies, and the inability to enforce existing mandates, summarised by one respondent as *”not all clubs or leagues have clear, enforced concussion management policies.”* **Limited access to qualified medical professionals** (B4; n = 25, 15.7%) was common and may have been driven by rural and regional respondents (Table 1). For example, *”in our rural area, availability of seeing a doctor can take weeks which delays any medical rehab.”* **Financial and resource constraints** (B6; n = 18, 11.3%) were noted by some respondents as prohibitive, particularly for families managing prolonged recovery requiring specialist input. For example, *”most parents struggle with financial costs of sport, and mild to severe concussions need proper management long term with physio, docs, test batteries, time off school.”* Finally, **player underreporting and symptom concealment** (B7; n = 11, 6.9%) was identified as a distinct barrier, with respondents noting that children are reluctant to disclose symptoms to avoid exclusion.

#### 3.3.3 Information and Resource Needs

Respondents were asked what they would like to know more about regarding concussion management. Of 151 responses received, 121 were substantive (30 excluded as non-substantive), and six categories were identified. **Education, training and club-level resources** (M5) was the most common area of desired knowledge (n = 47, 38.8%), with respondents seeking clearer protocols disseminated consistently across clubs, mandatory training for coaches and volunteers, and better access to information at the community level. As one respondent noted, *”management plans and guidelines available for all to see and not just online. If junior footy can somehow enforce time off as they do in the AFL.”* Another highlighted the need to have clear responsibilities described for each club-specific role, stating that *”the responsibilities of the coach and team when a child does receive a hit or knocks their head and how they respond to the situation.”* **Recognising symptoms and early identification** (M2) was the second most common category (n = 46, 38.0%), encompassing requests for guidance on identifying both typical and delayed presentations of concussion, distinguishing a concussion from *”just a knock,”* and knowing when to escalate to emergency care. The challenge of delayed concussion was also raised by respondents, with one stating *”my son had delayed concussion but it was difficult to diagnose so we erred on the side of caution.”*

The remaining four categories were identified by approximately one quarter or fewer of respondents. **Return-to-play protocols and recovery timelines** (M1; n = 31, 25.6%) and **home management and practical post-incident guidance** (M4; n = 31, 25.6%) were equally common, and encompassed what to do in the hours and days immediately after injury, and how to navigate safe return to sport. One respondent stated the need for a *”day by day timetable of what is acceptable at what point in post-concussion period. Saying ‘do nothing, just rest’ is not very helpful.”* **Long-term effects and ongoing monitoring** (M3; n = 29, 24.0%) reflected a desire for clearer evidence on cumulative risk, with respondents seeking both the latest research and information about what prolonged or repeated concussions mean for their child’s brain health. One respondent described the need to understand *”long-term effects, and how repeated concussions might affect a child’s brain and overall health.”* Finally, **equipment, technology and prevention** (M6; n = 19, 15.7%) was the least common category, with respondents raising questions about the effectiveness of helmets, the potential role of mouthguard technology and impact-tracking apps, and whether brain imaging is necessary.

### 3.4 Part B, Quantitative Data

#### 3.4.1 Subsample Characteristics

A total of 58 respondents (26.0% of the full sample) had a child who had experienced a concussion in the 12 months prior to completing the survey. Of these, a concussion had occurred within 1-4 months of completing the survey for the majority (n = 25, 43.1%), with smaller proportions having occurred 5-8 months (n = 18, 31.0%) or 9-12 months prior (n = 15, 25.9%). Concussions most often occurred during a match (n = 25, 43.1%) or in other contexts (n = 15, 25.9%), with smaller proportions occurring during training (n = 7, 12.1%) or a trial match (n = 3, 5.2%). Eight respondents (13.8%) did not report a location. Full information is presented in Table 5.

**Table 5:**
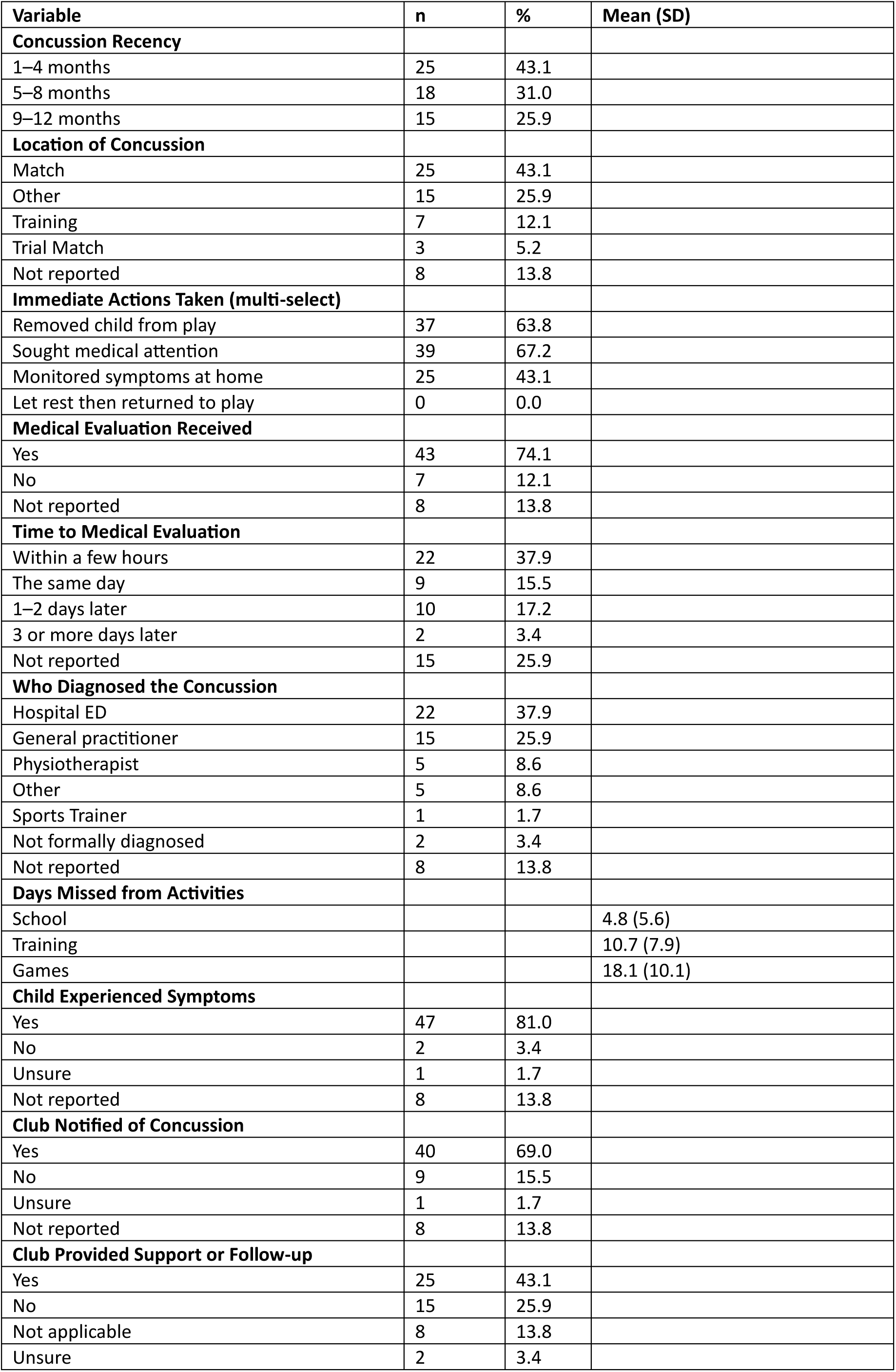
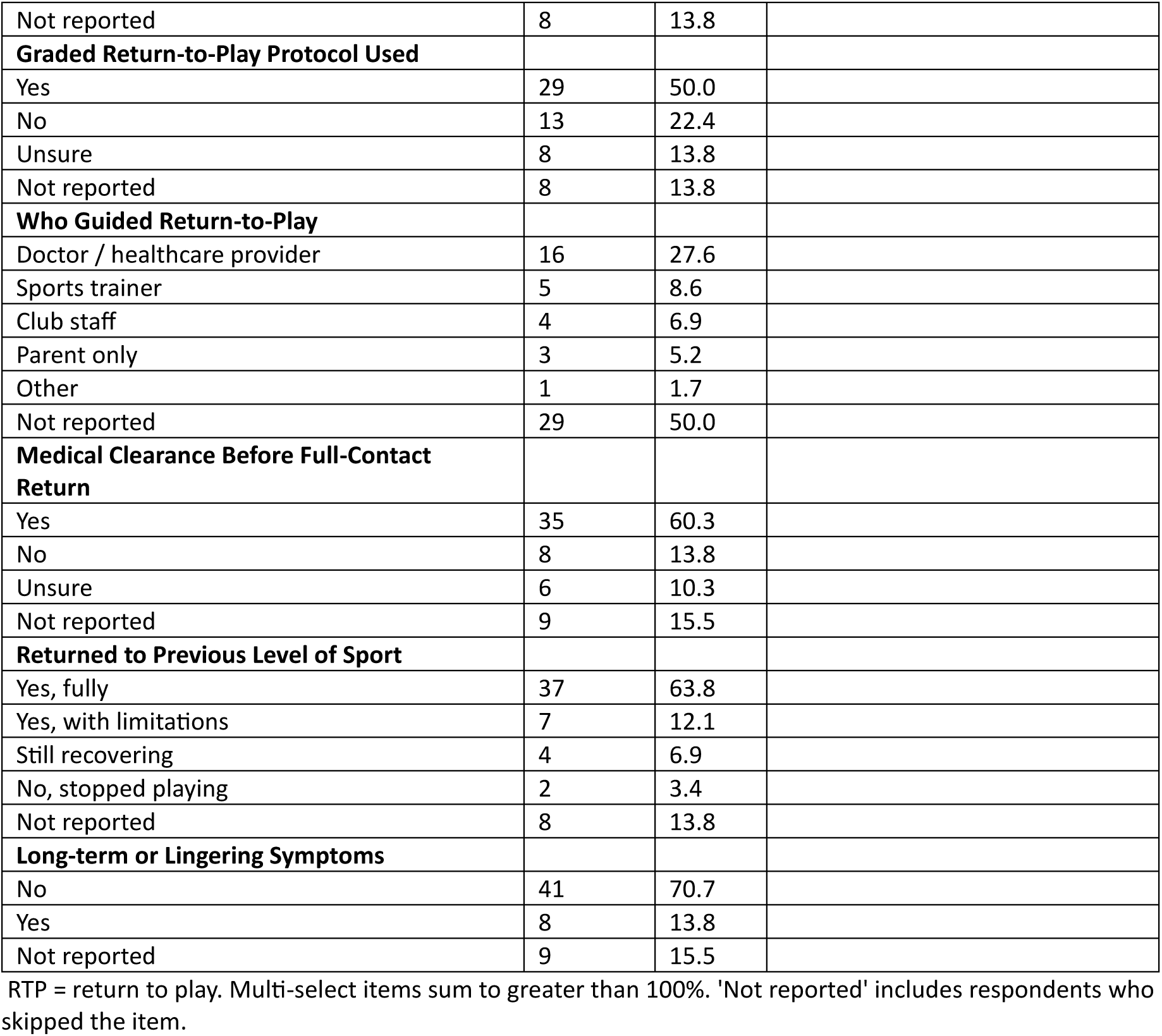
Concussion information for parents whose child/children had experienced a concussion in the last 12 months.

#### 3.4.2 Immediate Management

With respect to immediate actions taken following the concussion, 67.2% (n = 39) sought medical attention, 63.8% (n = 37) removed their child from play, and 43.1% (n = 25) monitored symptoms at home. No respondents reported allowing their child to rest and then return to play. Most children received a medical evaluation (n = 43, 74.1%), which most commonly occurred within a few hours (n = 22, 37.9%) or 1–2 days later (n = 10, 17.2%). Diagnosis was most often made by a hospital emergency department (n = 22, 37.9%) or a general practitioner (n = 15, 25.9%).

#### 3.4.3 Time Lost and Symptom Experience

The median number of days missed from school was 3.0 (M = 4.8, SD = 5.6, n = 45), from training was 12.0 (M = 10.7, SD = 7.9, n = 41), and games was 21.0 (M = 18.1, SD = 10.1, n = 40). Most children were reported to have experienced symptoms following their concussion (n = 47, 81.0%), though long-term or lingering symptoms were uncommon (n = 8, 13.8%).

#### 3.4.4 Return-to-Play Management and Club Support

Most respondents notified their club of the concussion (n = 40, 69.0%), and 43.1% (n = 25) reported receiving support or follow-up from the club during recovery. A graded return-to-play protocol was used in half (n = 29, 50.0%) of cases, which was often guided by a doctor or healthcare provider (n = 16, 27.6%). Medical clearance before returning to full-contact sport was obtained in 60.3% (n = 35) of cases, while 13.8% (n = 8) reported no medical clearance, and 10.3% (n = 6) were unsure. Most children returned to their previous level of sport (n = 37, 63.8%) or with some limitations (n = 7, 12.1%), with two children (3.4%) ceasing to play completely and four (6.9%) still recovering at the time of the survey.

### 3.5 Part B, Qualitative Data

#### 3.5.1 What worked well in the recovery process

Thirty-nine responses, of which 36 contained substantive content. Four categories were identified. **Rest and reduction of physical and cognitive load** (W1) was the most reported category (n = 26, 72.2%), with parents describing a combination of physical rest, removal of screens and devices, and time away from school as their approach. **Medical guidance and professional oversight** (W2; n = 12, 33.3%), **gradual structured return to activity** (W3; n = 8, 22.2%), and **family, school and social support** (W4; n = 8, 22.2%) were each identified by approximately one in five respondents. Notably, responses that spanned multiple categories tended to reflect more comprehensive management approaches. For example, one respondent stated, *”consistent medical guidance, rest, and gradual return-to-play monitoring helped ensure a smooth and safe recovery.”* This may suggest that positive recovery outcomes were associated not with any single strategy but with the integration of several concurrent approaches. All categories identified across open-ended questions in Part B are presented in Table 6.

**Table 6:**
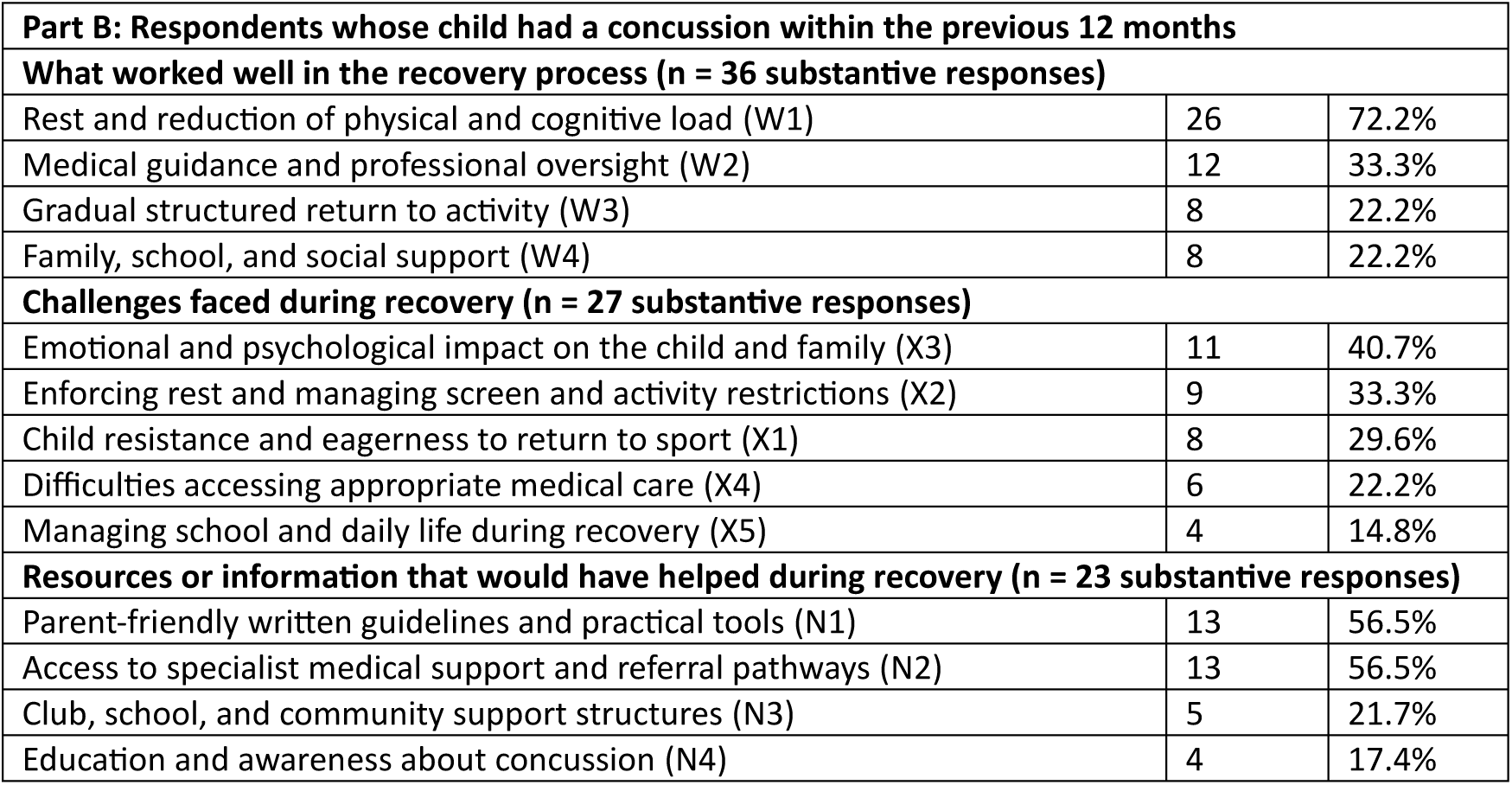
Summary of Part B qualitative analysis results.

#### 3.5.2 Challenges faced during recovery

Thirty-three responses were received for this question, with 27 included (six excluded as non-substantive). Five categories were identified. **Emotional and psychological impact on the child and family** (X3) was the most frequently cited challenge (n = 11, 40.7%), encompassing anxiety, boredom, frustration, behavioural changes, and parental worry. Respondents described children who were emotionally distressed by the experience, including reports of memory loss and behavioural change. For example, one parent stated that their child experienced *”memory loss, behavioural changes, agitated, emotional.”* Another described the psychological toll of the experience as a key challenge, *”emotional and mental state was just as bad as the injury.”* **Enforcing rest and managing screen and activity restrictions** (X2; n = 9, 33.3%) was the second most frequently identified challenge, reflecting the practical difficulty of keeping young individuals compliant with recovery protocols. Respondents described battles over screen time and physical inactivity. For example, *”getting him to rest, no screen time.”* **Child resistance and eagerness to return to sport** (X1; n = 8, 29.6%) captured the child’s excitement to return, rather than the practical issues challenged faced by the parent. Respondents described children who placed significant pressure on themselves to get back to their team. For example, *”once he was symptom free, the want to return too early was huge. We just had to get him to understand the repercussions of returning too soon.”* **Difficulties accessing appropriate medical care** (X4; n = 6, 22.2%) were reported by respondents. For example, *”access to medical facilities. We live in a rural town with a local hospital that often does not have a GP on site, distance from home location (we live on a farm)”*, alongside financial barriers including the cost of MRI and healthcare. Finally, **managing school and daily life during recovery** (X5; n = 4, 14.8%) was identified by a smaller subset of respondents, though several described this as compounding other challenges, with one parent stating *”managing schoolwork and daily routines while balancing rest and recovery was challenging, along with the initial worry about returning safely to sport.”*

#### 3.5.3 Resources that would have helped

Twenty-nine responses were received for this question, of which 23 were included (six were excluded), and four categories were identified. **Parent-friendly written guidelines and practical tools** (N1) and **access to specialist medical support and referral pathways** (N2) were identified by more than half of substantive respondents (n = 13, 56.5% each). Requests under N1 included simple written formats, such as *”maybe a booklet”*, *”kid friendly info”*, *”a concussion approved activity list,” as* well as more structured resources, such as *”clear, accessible guidelines”* and “*step-by-step instructions for parents on managing concussions at home.”* The frequency of N2 reflects a consistent finding across the Part B data that access to appropriately trained medical professionals was both highly valued and difficult to obtain, with respondents requesting *”closeness to a medical practitioner”* and *”access to a concussion specialist”* as their primary unmet need. One respondent captured both categories together, describing the ideal as *”clear written guidelines on concussion recovery, access to medical advice specific to young athletes, and more information about safe return-to-play timelines.”* **Club, school and community support structures** (N3; n = 5, 21.7%) and **education and awareness about concussion** (N4; n = 4, 17.4%) were identified less frequently, though several responses in N3 pointed to the value of proactive club-level communication *”perhaps info for coaches and trainers to hand to parents at time of injury would be helpful.”* One respondent in N4 highlighted the importance of equipping the child themselves with an understanding of their recovery: *”more child-friendly education so he understood why this was so important.”*

## 4. Discussion

This study assessed the concussion knowledge, attitudes, confidence, and management practices of parents and staff involved in junior community ARF, and identified key barriers and informational needs in this population. Collectively, the findings suggest that this population generally holds safe attitudes toward concussion management, but may be constrained by gaps in knowledge, structural support, and access to resources. These gaps manifest in low scores on sport-specific guideline knowledge, and in the real-world management experiences of parents whose children sustained a SRC, who identified that club support, graded return-to-play adherence, and access to appropriate medical care were inconsistent.

### 4.1 General concussion knowledge and aFtudes

Respondents demonstrated above average general knowledge of concussion, with a mean CKI score of 19 out of 25 (76.6%). These findings are consistent with prior reviews^17^ and primary studies^26^ suggesting that adults involved in youth sport tend to perform above average on general knowledge measures. However, two areas of knowledge deficit are noteworthy. Firstly, a proportion of respondents incorrectly believed that brain imaging typically shows visible damage following a concussion. Secondly, many respondents underestimated the risk of long-term harm from multiple concussions, despite evidence that repeated concussive injury is associated with cumulative consequences in children.^8^ These misconceptions are meaningful targets for educational intervention.

Attitudes were scored stronger than knowledge, with mean CAI scores corresponding to 86.8% of the maximum. This disparity, whereby respondents endorse broadly safe attitudes while holding partial or inaccurate knowledge, underscores the importance of targeting knowledge specifically in education programs rather than assuming that positive attitudes will translate to improved practice.^18^

### 4.2 AFL guideline knowledge

When responding to questions pertaining to the 2024 AFL Community Concussion Guidelines,^22^ respondents reported a mean score of approximately 9 out of 15 (60%). This may suggest that these guidelines are not being effectively communicated to the parents and staff responsible for their implementation, a finding that has been seen in prior research nationally^27^ and internationally.^28, 29^

It is important to note that this lack of knowledge was not always a cause for concern. For example, most respondents failed to identify the AFL-specified minimum monitoring period of at least three hours following a suspected concussion, instead selecting “until symptoms resolve”, which is more conservative. Nonetheless, these results broadly suggest that the guideline’s specific recommendations are not reaching end-users in a memorable or accessible format. In line with prior work,^30^ this reinforces the notion that the transition from guideline publication to embedded knowledge at a club level remains an ongoing challenge.

### 4.3 Confidence and knowledge

Despite moderate concussion confidence and good concussion knowledge at the cohort level, findings revealed no association between self-reported confidence and both general concussion knowledge (r = −.04) and attitudes (r = .09). This indicates that respondents’ subjective sense of how capable they are may not meaningfully reflect how much they know or how safely they consider concussion. Indeed, the only significant association with confidence was with AFL Guideline Knowledge (r = .33), suggesting that familiarity with sport-specific protocols may be a more meaningful driver of perceived competence than general knowledge alone. This may also suggest that people involved in junior ARF feel more confident in being able to manage concussion when they have specific guidelines and protocols to follow or refer to, rather than just relying on general knowledge alone. It is also possible that participants may have good theoretical knowledge surrounding concussion, but simply lack the confidence to apply that practice in real-world situations, which would be expected when considering the population sampled were not trained in a medical context.

This finding also has implications for how concussion education programs are designed and evaluated. Self-reported confidence is frequently used as an outcome measure in educational interventions, partially because it is easy to assess.^17^ However, if confidence does not track with knowledge, then improvements in confidence scores are alone unlikely sufficient evidence that education programs are working. Future education programs in community ARF should incorporate objective knowledge assessments, rather than relying on self-reported confidence as a primary outcome. This also reinforces the need for ARF clubs to provide greater structural support and resource accessibility surrounding concussion management to parents and staff, to improve knowledge translation.

### 4.4 Concussion management

The present findings also provide a mixed picture of current concussion management. Positively, no respondents reported allowing their child to rest and return to play in the same match, 74.1% sought medical evaluation, and 60.3% obtained medical clearance before returning to sport. These figures compare positively with the findings of Haran et al. (2016),^19^ who reported that 42% of concussed children in community ARF were not managed according to recommended guidelines. This may suggest that some improvements in immediate management practices may have occurred over the past decade, indicative of greater concussion awareness, knowledge, and attitudes. Similarly, the median number of days missed from games (21 days) is consistent with the AFL’s updated minimum 21-day return-to-play guideline.

However, some findings warrant concern. In those whose child had experienced a concussion in the previous 12 months, a graded return-to-play protocol was used in only half of cases, and club follow-up and support during recovery was received by less than half of respondents (43.1%). Non-adherence to graded return-to-play protocols has been identified as a significant problem in grassroots contact sports,^31^ and the present findings suggest this remains an area requiring targeted support. This is noteworthy as the AFL community concussion guidelines clearly recommend that junior football nominate a concussion officer to oversee the management of young players with concussion or a suspected concussion, further suggesting that they are not being disseminated effectively at the junior level.

### 4.5 Barriers to concussion management

Knowledge and education gaps were identified as one of the primary barriers to concussion management. Importantly, respondents did not frame this as a personal knowledge deficit, but as a systemic problem affecting coaches, parents, players, and volunteer staff. This is noteworthy, as it suggests knowledge deficits could be considered structural problems requiring structural solutions, such as mandated and consistently delivered education rather than optional training. This speaks to need to build support within clubs, shifting attitudes and norms and creating more supportive club culture around concussion management.

Almost one third of respondents identified persistent “old school” or “she’ll be right” cultural attitudes as a barrier, a finding that has been documented across multiple sports.^16, 32^ This coincided with the category of player underreporting across both the concerns and barriers domains. This may suggest that parents are concerned that their children might conceal symptoms to remain on the field, and that club culture may also be a barrier to safe management. Addressing is likely to require not just individual education, but also changes at the club and league level to encourage the widespread social support of safe management practices.^33^

### 4.6 Structural and resourcing barriers

Structural barriers, including reliance on volunteers, absence of clear enforceable protocols, limited access to qualified medical professionals, and financial constraints, were identified by a substantial proportion of respondents. The barrier of limited medical care is likely reflective of the rural and regional respondents, consistent with evidence that rural communities face the longest delays to specialist concussion care.^34^ This has direct implications for the design of concussion management resources. For example, protocols and decision-support tools may need to function in settings where a physician or sports trainer is not present, where the person making decisions may have limited training.

### 4.7 Information and resource needs

The qualitative findings from Part B suggest that parents want accessible, practical, and specific information. The two most requested knowledge areas were education and club-level resources, and recognising symptoms and early identification, followed closely by return-to-play protocols and home management guidance (each commented on by approximately one quarter of respondents).

Collectively, these categories describe a desire for practical and clear guidance on symptom recognition, immediate management, recovery support, and return-to-play. Considering that the previously described AFL concussion resources are publicly available reinforces the suggestion that awareness and accessibility, rather than content, may be the limiting factors. One respondent’s observation that guidelines should be “available for all to see and not just online” captures a broader accessibility problem, whereby digital resources may not reach the parents and volunteers who are most likely to be making decisions in real time.

### 4.8 Recommendations for leagues and clubs

The following recommendations are made across three implementation levels, being governing bodies and leagues, individual clubs, and parent and family support.

#### 4.8.1. Governing bodies and leagues

1. Mandate and verify concussion education across all volunteer roles, not only coaches. The present findings suggest that education should also target managers, trainers, committee members, and parents.
2. Translate guidelines into brief, format-appropriate resources for non-clinical audiences.
3. should consider complementing current guidelines with a brief visual summary or quick-reference card covering immediate actions, the minimum monitoring period, and the return-to-play timeline.
4. Reinforce the importance of safe removal from play. Given the presence of cultural barriers to reporting, and the concern about player symptom concealment, leagues should consider campaigns that reframe removing a player from play as an act of good coaching and good sportsmanship.
5. Ensure consistent enforcement of return-to-play requirements. Only 50% of Part B respondents used a graded return-to-play protocol, and only 43.1% received club support during recovery. Leagues should implement mechanisms for tracking and supporting return-to-play progressions.

#### 4.8.2 Individual clubs

1. In line with current AFL concussion guidelines, clubs should designate one or multiple concussion officers who are trained in the relevant concussion protocols, and who are accountable for follow-up. Their role could include notifying relevant parties, support return-to-play progression, and maintaining contact with the family throughout recovery.
2. Provide written concussion action plans to all parents at the point of registration. Multiple Part B respondents indicated that they would have benefited from receiving clearer written information at the time of injury. Providing a concussion information sheet as part of the registration process of during pre-season process help ensure that parents have access to guidance before they need it.
3. Ensure trained personnel have clear decision-making authority. Some respondents noted that volunteer trainers and first aid officers often face pressure from coaches, players, and parents when making removal-from-play decisions. Clubs should have explicit, well-communicated policies affirming that the first aid officer or designated concussion officer has authority to remove a player from play, regardless of the score or the significance of the match.

#### 4.8.3 Supporting parents and families

1. Develop and widely disseminate a practical day-by-day home recovery guide. Resources such as a day-by-day recovery checklist, screen time guidance, and a simple symptom monitoring log would be a practical tool that parents could use during the recovery process.
2. Proactively signpost parents to appropriate medical referral pathways. Where local specialist care is unavailable (i.e., regional and rural communities), telehealth concussion services and primary care referral pathways should be signposted by clubs at the time of injury, including identification of public and private clinics that provide thorough multimodal post-concussion assessments.

### 4.9 Strengths and limitations

This study has several strengths. It is the first to examine concussion knowledge, attitudes, and management practices in junior community ARF since the AFL introduced updated community guidelines in 2024. The mixed-methods design, incorporating both validated measures and qualitative analysis, allowed a rich characterisation of the barriers and needs of this population. The use of rigorous data quality procedures, including IP geolocation, strengthens confidence in the authenticity of the data.

The limitations of the study should be acknowledged. The sample was self-selected and recruited primarily through community sporting networks and social media, which may have introduced systematic biases. For example, toward respondents with pre-existing interest in concussion management or with higher levels of health literacy. Accordingly, the findings may overestimate knowledge and awareness levels in the broader population of junior ARF parents and staff. The sample was also predominantly female (67.3%), which may limit generalisability to male caregivers and coaches.

## 5. Conclusions

Parents and staff involved in junior community ARF generally hold safe attitudes toward concussion management but lack the confidence, knowledge, structural supports, and accessible resources required to translate those attitudes into safe practice. The barriers to better practice are simultaneously educational, cultural, structural, and financial, and are unlikely to be resolved by any single intervention. The recommendations offered in this paper are intended to be practically implementable within the existing framework of the AFL’s community concussion recommendations and are grounded in the expressed needs of the community stakeholders who participated in this research. Addressing the translation gap between guideline publication and embedded community practice represents the most pressing priority.

## Data Availability

For transparency, all de-identified quantitative and qualitative data are available on the Open Science Framework project linked to this study (https://osf.io/cwdu4/files/osfstorage).

